# Automatic diagnostic support for diagnosis of pulmonary fibrosis

**DOI:** 10.1101/2024.08.14.24312012

**Authors:** Ravi Pal, Anna Barney, Giacomo Sgalla, Simon L. F. Walsh, Nicola Sverzellati, Sophie Fletcher, Stefania Cerri, Maxime Cannesson, Luca Richeldi

## Abstract

Patients with pulmonary fibrosis (PF) often experience long waits before getting a correct diagnosis, and this delay in reaching specialized care is associated with increased mortality, regardless of the severity of the disease. Early diagnosis and timely treatment of PF can potentially extend life expectancy and maintain a better quality of life. Crackles present in the recorded lung sounds may be crucial for the early diagnosis of PF. This paper describes an automated system for differentiating lung sounds related to PF from other pathological lung conditions using the average number of crackles per breath cycle (NOC/BC). The system is divided into four main parts: (1) preprocessing, (2) separation of crackles from normal breath sounds, (3) crackle verification and counting, and (4) estimating NOC/BC. The system was tested on a dataset consisting of 48 (24 fibrotic and 24 non-fibrotic) subjects and the results were compared with an assessment by two expert respiratory physicians. The set of HRCT images, reviewed by two expert radiologists for the presence or absence of pulmonary fibrosis, was used as the ground truth for evaluating the PF and non-PF classification performance of the system. The overall performance of the automatic classifier based on receiver operating curve-derived cut-off value for average NOC/BC of 18.65 (AUC=0.845, 95 % CI 0.739-0.952, p<0.001; sensitivity=91.7 %; specificity=59.3 %) compares favorably with the averaged performance of the physicians (sensitivity=83.3 %; specificity=56.25 %). Although radiological assessment should remain the gold standard for diagnosis of fibrotic interstitial lung disease, the automatic classification system has strong potential for diagnostic support, especially in assisting general practitioners in the auscultatory assessment of lung sounds to prompt further diagnostic work up of patients with suspect of interstitial lung disease.

## 1. Introduction

Idiopathic pulmonary fibrosis (IPF) is the most common and the most severe of fibrotic interstitial lung diseases (ILDs) [1], with a median survival time of only 3 years [2-4]. Apart from lung transplantation at a very early stage of the disease [5], no curative treatment is currently available, and treatment is focused on quality and prolonging of life [6]. The primary symptoms and signs of the condition include dyspnea, cough, inspiratory crackles, and finger clubbing [7,8]. It has been shown that delayed access to tertiary health care is associated with a higher mortality rate in IPF regardless of disease severity [9]. Several factors can contribute to delays in referring patients with suspected IPF to a tertiary health care. For instance, patients might experience symptoms such as cough and exertional dyspnea for months or even years before seeking consultation from their primary care physician. Additionally, due to the rarity of IPF, primary care physicians may not immediately recognize the condition, and key clinical signs, such as Velcro-like crackles during lung auscultation, may be missed [6].

Although IPF is considered a rare condition, it accounts for 17% to 37% of all ILDs. The impact of IPF is substantial, affecting social, healthcare, and economic aspects [10]. Given the disease’s fatal nature, it is crucial to achieve an accurate and timely diagnosis [7]. Recently, the first pharmacological treatments able to slow the progression of this disease have been developed [3], [11]. Therefore, diagnosing IPF at the earliest possible stage has become even more important.

Lung sounds can provide useful information regarding type and severity of pulmonary disease. In IPF, a common indicator, often used informally during lung auscultation by clinicians is the presence of fine crackles, often referred to as Velcro crackles, and described as high-pitched, impulsive sounds occurring mostly during inspiration. Recently, Sgalla et al. [12] showed that Velcro crackles in recorded lung sounds may facilitate early detection of fibrotic lung disease. However, identifying Velcro crackles can be difficult, especially for physicians under poor listening conditions, mainly due to the transient non-stationary characteristics of these sounds [13]. Furthermore, individual auditory acuity due to differences in training, experience and sensitivity of hearing often result in inter-examiner disagreement about the presence of crackles, potentially resulting in misdiagnosis and even mistreatment. Therefore, at present the gold-standard diagnosis for fibrotic lung diseases remains expert evaluation of high-resolution computed tomography (HRCT) scans of the lungs.

In the early course of IPF, crackles are typically generated in the base of the lungs, where the fibrosing process begins, but, as the disease progresses, crackles can also be heard in upper zones [14]. The number of crackles per breath cycle (NOC/BC) is related to disease severity in interstitial lung diseases [15]. Therefore, the aim of this study was to explore whether an automatic system based on average NOC/BC calculated from lung sounds recorded from the posterior lung bases, performs as well as assessment by a clinical expert in terms of classifying subjects as either having: pulmonary fibrosis, or not. Such a method, in conjunction with other assessments, might be used as a supporting tool in clinics for diagnosis of pulmonary fibrosis.

In this study, outcomes from an automatic classification method are presented, and validated against a gold standard diagnosis given by two radiologists obtained from HRCT images recorded on a dataset of 48 subjects. The capability of the automatic method to indicate presence of pulmonary fibrosis is also compared with that of auscultation by two expert respiratory physicians.

The methodology consists of pre-processing of the recorded lung sounds, automatic separation of crackles from normal breath sounds, crackle verification and counting, estimation of NOC/BC, and exploration of the ability of the average NOC/BC to classify pulmonary fibrosis subjects and non-pulmonary fibrosis subjects. The study represents the first steps towards development of an automatic classification system based on the average NOC/BC estimated using lung sounds recorded from the posterior lung.

## 2. Data description

Subjects for this study were recruited in the Radiology Units of the University Hospitals of Modena in Italy. Participants were recruited from among those referred to the clinics for an HRCT scan of the lungs.

The study was approved by the local ethics committee of Modena, Italy. Written informed consent was collected from all participants. For those who agreed to take part in the study, demographic information, medical history and family history of ILD were collected, together with the clinical indication for performing the HRCT scan (prescription slip or referral letter from general practitioner or respiratory physician). Lung sounds were then recorded as detailed below. Following these assessments, the subjects underwent the HRCT scan of the chest.

For each subject, the lung sounds were recorded using an electronic stethoscope (Littmann 3200, 3M, USA) from six different sites on the posterior chest – four recordings were taken at the lung bases (two at left and two at right side) and two recordings at mid chest (one per side). The recordings were sampled at 4 kHz with a resolution of 16-bits. Subjects were asked to breathe deeply, and the stethoscope was kept as still as possible through the recording process to reduce background noise. After each recording, a small, radio-opaque metallic mark (a bio-compatible electrocardiography electrode) was applied to the skin to allow visualization and hence correlation of the recording sites on the HRCT. The recordings were transferred via a Bluetooth connection to the computer (using the Littmann StethAssist software) and saved in the .wav format for further analysis. However, for the analysis each lung sound recording was up-sampled by a factor of 11.025, increasing the sampling frequency to 44, 100 Hz. The sampling rate of 44,100 Hz was selected because it is recommended by Cheetham et al. [16] for respiratory sound recordings. Recordings at different sites were consecutive, not simultaneous. In this study, only lung sounds recorded from the lung bases: (as shown in Fig. 1 with green text: L2, L3, L5 and L6) were used for further analysis since, as mentioned in [14], at the early stage of IPF crackles are typically heard only in the lower lung.

**Fig. 1.**
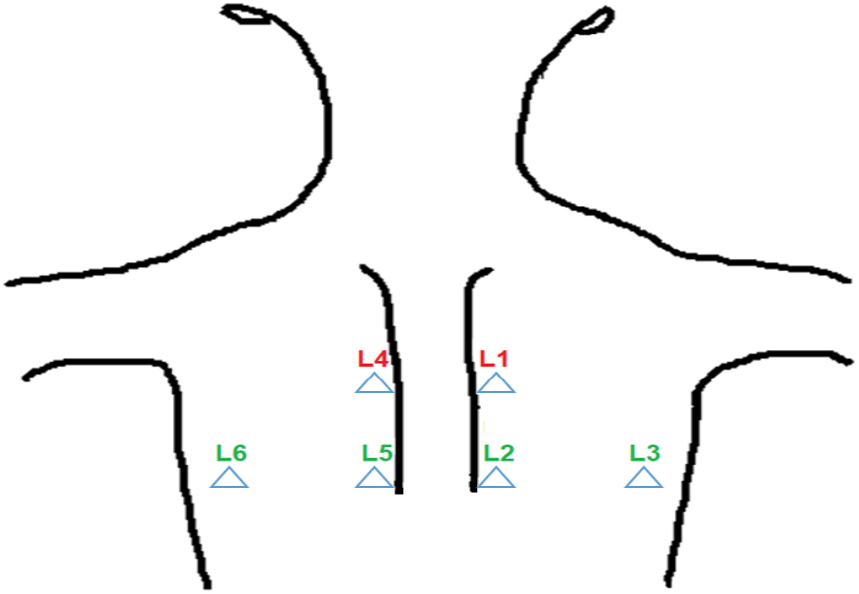
Lung sounds recording sites (L1-L6), in this study lung sounds recorded from 4 posterior base locations (L2, L3, L5 and L6, indicated in green font) are analyzed.

The single HRCT sections corresponding to the visible marked sites of recording were extracted for each study participant, randomized and independently reviewed by two expert radiologists who noted the presence or absence of pulmonary fibrosis. In the case of disagreement, fibrosis was marked as present. The marked images were used as the ground truth for this study with a subject labelled as ‘pulmonary fibrosis’ if any single image slice was so labelled. The dataset consisted of 185 lung sound files, recorded from 48 subjects, of which 24 subjects had been diagnosed from the HRCT as fibrotic and 24 age and gender-matched subjects diagnosed as not fibrotic (Table 1). Note that the not fibrotic subjects had all been referred to the clinic for a CT scan and therefore did have symptoms indicating potential presence of a pulmonary disease of some kind.

**Table 1.** Demographic data and diagnosis.

|  |  | **Fibrotic**<br>**(No. of**<br>**subjects=24)** | **Not fibrotic**<br>**(No. of**<br>**subjects=24)** |
| --- | --- | --- | --- |
| **Mean(sd) Age, years** |  | 69.83 (9.24) | 69.79 (10.54) |
| **Sex (%)** | Male | 15 (62.5 %) | 15 (62.5 %) |
|  | Female | 9 (37.5 %) | 9 (37.5 %) |

The sound files were also assessed by two expert respiratory physicians with expertise in ILD, one working at the University Hospital of Southampton, UK and one at the University Hospital of Modena, Italy. They were blind to the HRCT assessment of the images and were asked to listen to the sound files played, using Audacity software, through identical sets of headphones (Sennheiser HD201 closed dynamic stereo) and to rate them for the presence or absence of “Velcro-type” crackles. In cases of disagreement, a file was marked as Velcro crackles. The sound files were presented in random order to the two raters to reduce subjective effects due to e.g. fatigue and to ensure that files from the same subject were not clustered together.

## 3. Steps used for estimating number of crackles per breath cycle

The number of breath cycles in each lung sound file was audio-visually marked in Audacity by an experienced researcher, and only full breathing cycles were used for the analysis. The steps used for estimating NOC/BC are shown in Fig. 2 and described below.

**Fig. 2.**
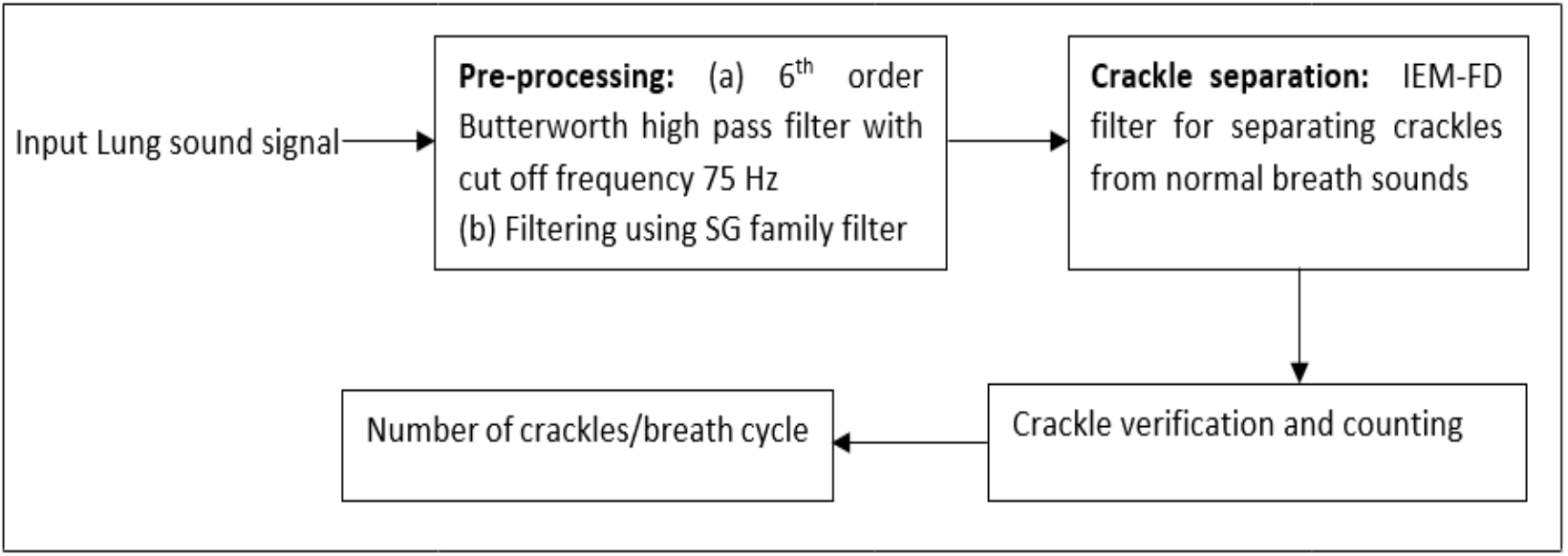
Steps used for estimating number of crackles per breath cycle.

### 3.1. Preprocessing

Firstly, each lung sound signal was filtered using a 6^-th^ order Butterworth high-pass filter with cut-off frequency of 75 Hz for eliminating the low frequency components related to movement artifacts, heart sound, muscle artifacts etc. Secondly the signal was smoothed using a filter from the Savitsky-Golay (SG) family to eliminate the high frequency, low amplitude ripples in the signal. The values of the SG filter parameters are: degree of fitting polynomial *p*_*f*_ = 4 and number of coefficients *n*_*c*_ = 89 [17]. Figs. 3 (a) and (b) show the input lung sound signal and the pre-processed lung sound signal, respectively.

**Fig. 3.**
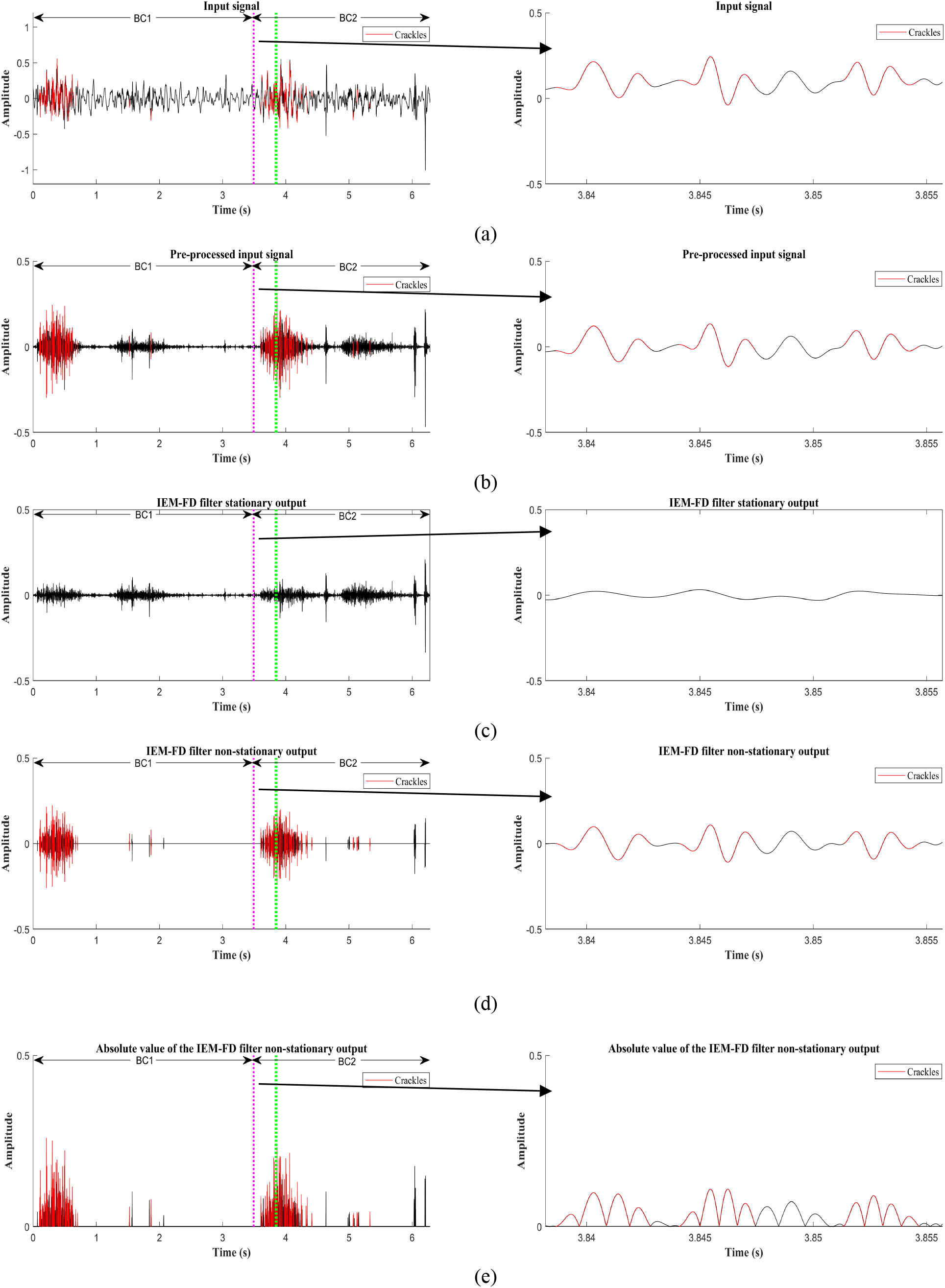
Estimation of NOC/BC. (a) Input lung sound signal. (b) Pre-processed input lung sound signal. (c) IEM-FD filter stationary output. (d) IEM-FD filter non-stationary output. (e) Absolute value of the non-stationary output of the IEM-FD filter (BC1- first breath cycle, BC2- second breath cycle). Figures to the right in each sub-plot show an expanded view of the section marked by the vertical pink and green dotted lines. Red-coloured areas of the plots indicate places where crackles have been identified by the analysis process described in the text.

### 3.2. Crackle separation

Separation of crackles from background normal breath sounds can lead to better estimation of the number of crackles present. Therefore, after preprocessing, an automatic crackle separation technique, the iterative envelope mean fractal dimension (IEM-FD) filter [17], was used to separate crackles from normal breath sounds. The IEM-FD filter was selected for crackle separation due to its high accuracy for identifying both fine and coarse crackles, ability to preserve crackle morphology after separation, and its low computational cost [17]. The IEM-FD filter combines two processes: the IEM method and the FD technique [18]. The IEM method estimates the non-stationary and stationary parts of the lung sound signal. The time duration of each crackle is typically less than 20 ms and therefore they can be considered as non-stationary compared to the timescale of the quasi-stationary normal breath sounds hence the IEM filter sends the majority of crackles to its non-stationary output, whereas the stationary output mainly consists of the normal breath sounds. The FD technique is then applied to the non-stationary IEM output for further refinement of the separation process. Figs. 3(c) and 3(d) respectively show estimates from the IEM-FD of the stationary and non-stationary outputs of a lung sound signal.

### 3.3. Crackle verification and counting

An ideal crackle separation technique should separate only crackles into one estimate and only normal breath sounds into the other. However, in practice some part of the normal breath sounds may remain in the estimate of the crackle signal. Therefore, after the initial separation process, each potential crackle was verified to ensure it meets a prescribed set of rules defined by Murphy et al., [19], Pinho et al., [20], and additional conditions empirically generated from the data [21].

In detail the process to verify each crackle is as follows:

- The absolute value of the non-stationary signal (ANST) is first calculated (as shown in Fig. 3 (e)).
- The two-cycle deflection width (2CD) of a crackle is the duration of its first two full cycles of oscillation. These first two oscillation cycles correspond to 5 consecutive peaks in the ANST. To search for potential crackles, a window just long enough to include the first 6 peaks in the ANST is defined. Including 6 peaks guarantees that the 2CD of any crackle starting just after the first peak will be inside the window. If all 5 valleys in the ANST correspond to zero crossings of the crackle signal (see Fig. 4), that window is considered as a potential crackle window (PCW) and is explored further. The valley after the first peak of the PCW is considered to be the starting point of a potential crackle (PC). On the other hand, if there is no correspondence between the peaks of the ANST and valleys in the crackle signal, no further exploration of that window occurs and instead, a new window containing 6 peaks starting from the second peak of the original window is generated. The process continues with the variable length window moving along by one peak each time until a PC is identified. Note that if the lung sound signal is not smoothed by the SG filter in the pre-processing step the crackle signal and the ANST may contain additional high-frequency peaks and valleys not associated with the crackle 2CD.
- When a PCW is identified, two further windows are defined: the before window (BW) and the after window (AW). As shown in Fig. 5 the BW contains 5 valleys just before the PCW including the first valley of the PCW, and the AW is similarly constructed, containing the last valley of the PCW. Note if there are fewer than 5 valleys in the signal before the PCW, the BW is not defined (see C1 in Fig. 5) and, correspondingly, the AW is not defined if there are fewer than 5 valleys after the PCW. The choice of 5 valleys for the BW and AW is to keep their length as close as possible to the length of the PCW in order to minimize identification of false crackles but maximise identification of true crackles. With a longer BW and AW, true crackles may be missed and for a shorter BW and AW false crackle detection may increase.
- Next each PC is verified using the following criteria:
  1. The beginning of the event has a sharp deflection in either a negative or a positive direction [19, 22].
  2. After the initial deflection width (IDW), the time between baseline crossings of successive peaks is progressively longer [19, 22]. However, Pinho et al., [20], note that not all crackles follow the standard rules, therefore ± 50 % deviation of time between peaks is selected to verify this condition [21].
  3. The IDW of the potential crackle is at least 8 times smaller than the largest deflection width (LDW) [20].
  4. The amplitude of the LDW peak of the crackle is greater than all other peaks of the crackle [21].
  5. The PCW mean is greater than 1.2 times the BW mean [21].
  6. The PCW mean is greater than the AW mean. Note that conditions (4) and (5) not only help to count temporally overlapping crackles but also ensure that any crackle with more than 5 zero crossings is not considered more than once [21].
  7. The amplitude of the IDW peak and LDW peak of the potential crackle are greater than one peak before the IDW peak (amplitude of the first peak of the PCW) [21].
  8. The 2CD of the PC is less than 20 ms [21].
  9. The IDW of the PC is less than 3 ms [21].
- If a PC fails to meet one or more of the above conditions, it is not counted as a true crackle and the next window of 6 peaks is defined starting from the 2nd peak of the existing PCW. On the other hand, if a PC meet all the conditions, it is marked as a true crackle. The new window then starts from the first peak after the existing PCW. In every case the window length varies to ensure 6 peaks are contained within.
- All validated crackles are then considered to be true crackles.

**Fig. 4.**
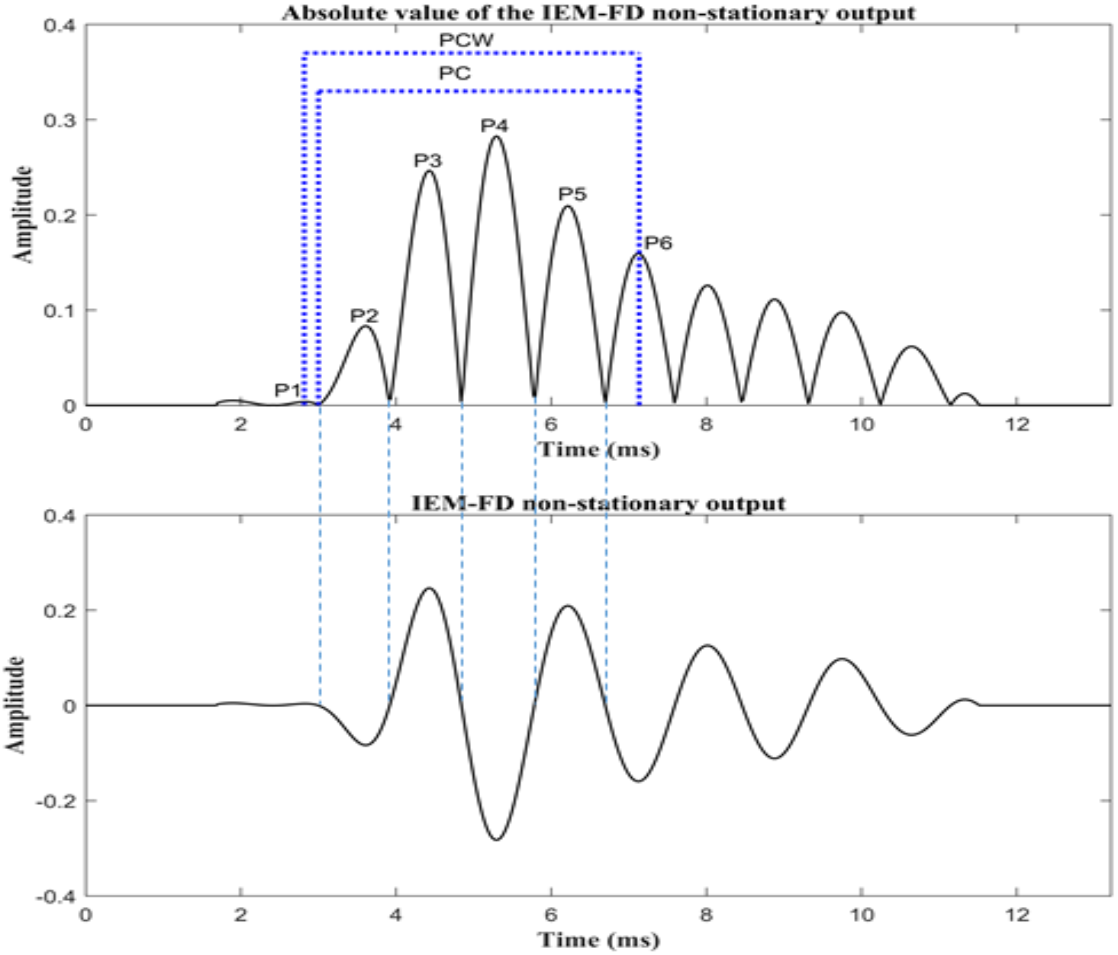
A potential crackle window (PCW) with potential crackle (PC) and six estimated peaks (P1-P6), where it is shown that all the valleys between the six peaks of the PCW correspond to the zero crossings of the IEM-FD filter non-stationary output.

**Fig. 5.**
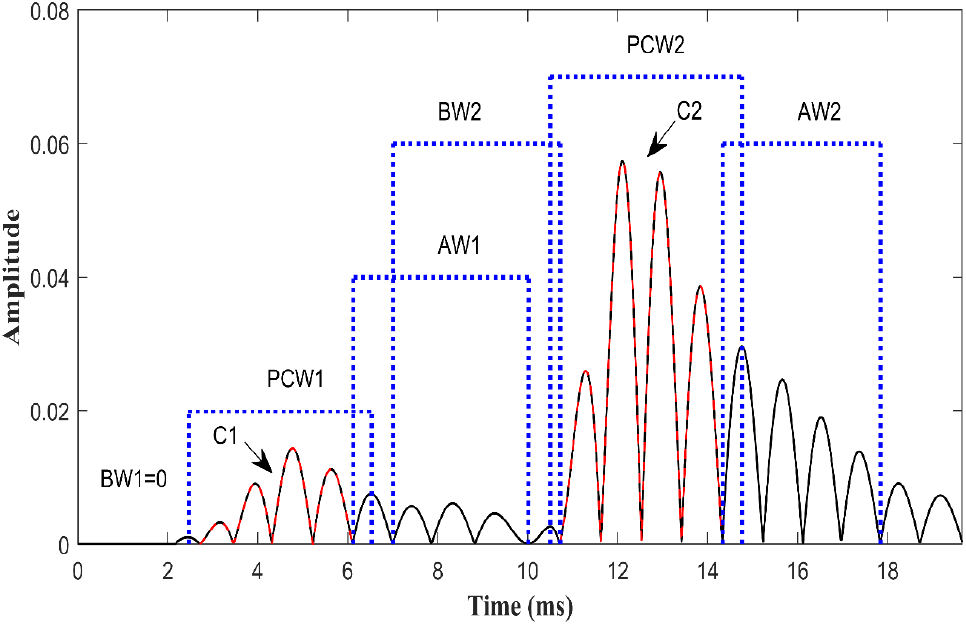
Absolute value of the crackle signal containing two crackles C1 and C2 with their respective potential crackle windows (PCW1 and PCW2), before windows (BW1 and BW2) and after windows (AW1 and AW2).

To aid visualization of the automatically detected crackles, a section of 0.018 s of the second breath cycle (BC2 in Fig. 3 is shown separately in the right side of Fig. 3 (a – e). Note that the crackle detection method is not assessed in terms of sensitivity or positive predictive value because of the lack of a reference signal to compare with. Therefore, it is possible some crackles in the signal are missed and that some false crackles are detected. This could be due to true crackles occurring which do not meet established crackle verification conditions or because the IEM-FD filter fails to separate all the crackles from the input lung sound signal. However, the process of separation using the IEM-FD filter has previously been shown to perform with high accuracy (more than 99 % of fine and coarse crackles) in directing true crackles to its non-stationary output and minimising the inclusion of contributions from the normal lung sounds [17]. Further, many of the crackle verification conditions used here have been tested in previous studies [19, 20, 22].

### 3.4. Number of crackles per breath cycle

Once all true crackles were verified, the average number of crackles per breathing cycle (NOC/BC) was calculated for each subject. Table 2 shows the average NOC/BC calculated for fibrotic and non-fibrotic groups. The difference in average NOC/BC between the two groups was significantly different (*t* =4.94, p<0.001) under an independent sample t-test.

**Table 2.** Average NOC/BC for fibrotic and non-fibrotic groups. Data are presented as the mean (standard deviation).

| Parameter | Fibrotic (No. of subjects=24) | Non-fibrotic (No. of subjects=24) | p-value |
| --- | --- | --- | --- |
| Average NOC/BC | 31.47 (11.82) | 16.43 (9.11) | <0.001 |

## 4. Receiver operating characteristic (ROC) curve

A receiver operating characteristic (ROC) curve was generated (see Fig. 6) for the average NOC/BC to determine the potential of the average NOC/BC to differentiate the two groups of subjects (fibrotic or non-fibrotic). The average over all recordings of NOC/BC = 18.62 was taken as a cut-off (AUC=0.845, 95 % CI 0.739-0.952, p<0.001; sensitivity=91.7; specificity=59.3) to differentiate the two groups based on its performance in terms of sensitivity and specificity. The crackles in IPF patients are usually generated in the inspiratory phase. Flietstra et al. [23] found that the mean number of crackles in the inspiratory phase in IPF patients was 18, which further supports the selection of 18.62 as a cut-off for the average NOC/BC.

**Fig. 6.**
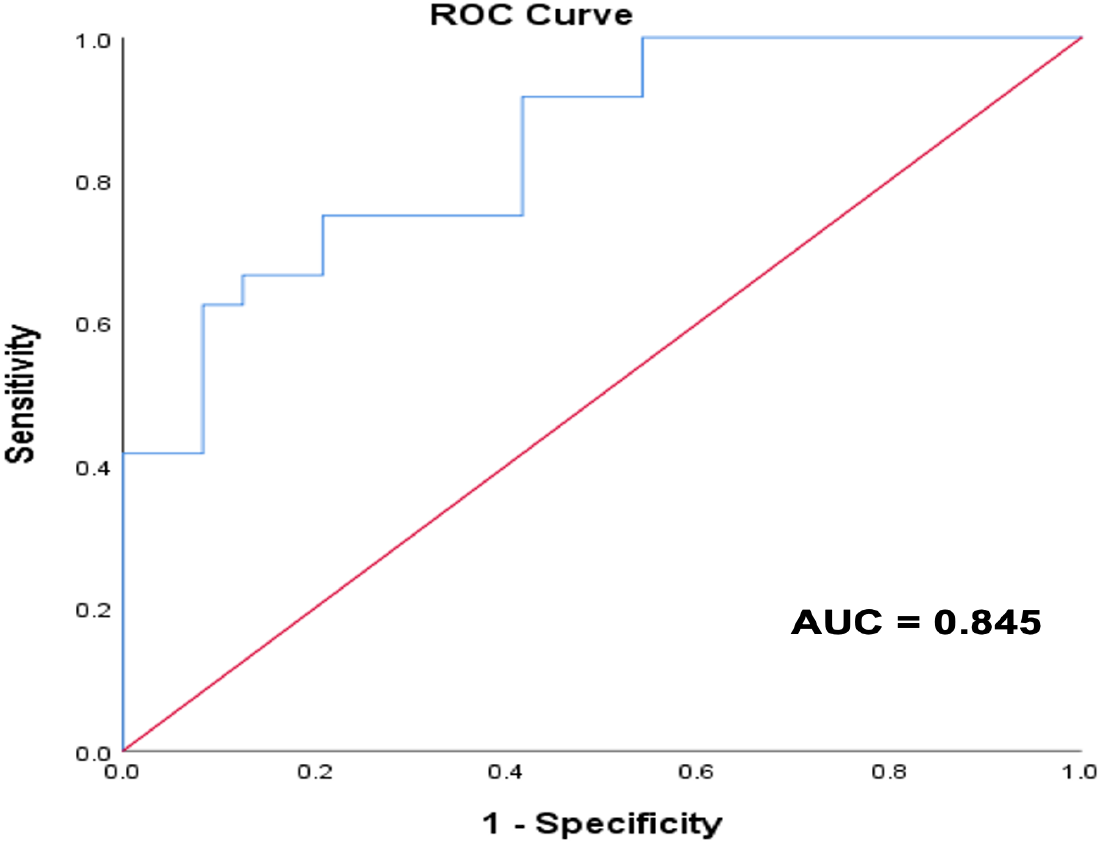
Receiver operating characteristic (ROC) curve for the average NOC/BC.

## 5. Assessment by physicians

Two expert physicians independently listened to each recoded lung sound file presented by Audacity software through over-ear-headphones (Sennheiser HD201 closed dynamic stereo) and marked each recording for presence or absence of Velcro crackles. In case of disagreement, a recording was marked as Velcro crackles present. Where at least one physician identified Velcro crackles as present in at least one recording site for a given lung base (left or right) that lobe was identified as having Velcro crackles present. If at least one lung base had Velcro crackles, the subject was classified as having Velcro crackles. Subjects with no Velcro crackles identified in either lobe were placed in the no Velcro crackles category. The physician assessment was compared to the evidence from the HRCT scans and to the classifications from the automated system.

## 6. Results and discussion

The classification performance achieved by each of the two experienced physicians, their combined (average) performance, and the performance of the automatic system are presented in Table 3.

**Table 3.**
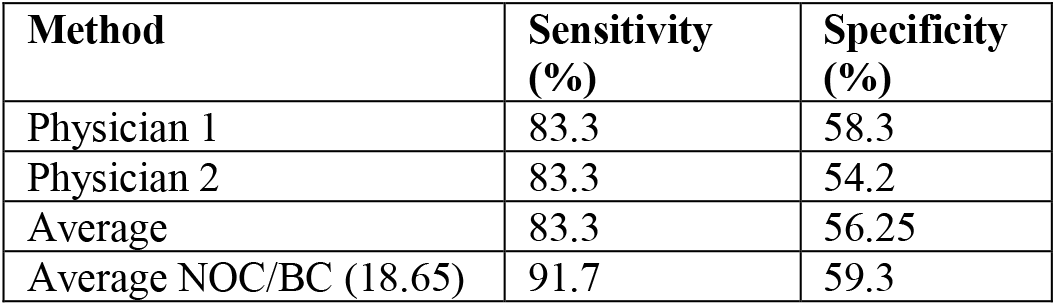
Classification performance of the physicians and the automatic system.

The two physicians had equal sensitivity (83.3 %), which showed their ability to identify Velcro crackles in subjects diagnosed with fibrotic lung disease, was high. However, their average specificity, at 56.25 % was lower and specificity was similarly lower for each physician individually.

The automated system using the average NOC/BC with an 18.65 cut-off value achieved a higher sensitivity (91.7 %) than either physician individually or on average and a similar specificity (59.3 %) to the higher of the two physician individual scores. Further, the automatic system identified more than 90 % of the same subjects as having fibrosis as the physicians which suggests the diagnostic match between the performance of physicians and the automatic system is good.

Crackles can play a crucial role in early diagnosis [24,25]. Notably, Velcro crackles have been suggested as a key indicator of early diagnosis of IPF [26, 27]. Several studies have explored their use in classifying patients and diagnosing lung diseases. For instance, Flietstra et al. [23] demonstrated that the characteristics of crackles in IPF may help distinguish this condition from congestive heart failure and pneumonia. Additionally, Baughman et al. [28] noted that the presence of crackles can aid clinicians in differentiating sarcoidosis from IPF. Moreover, Melbye et al. [29] evaluated the utility of crackle characteristics in diagnosing chronic obstructive pulmonary disease (COPD) and found that early inspiratory crackles are strong predictors of the condition.

Lung sounds have been very useful in medicine for checking respiratory health or disease, especially since the stethoscope was invented [30]. Recorded lung sounds are particularly instrumental in classifying pulmonary pathologies [31,32]. There has been a significant advancement of lung sounds recording devices over the years [33,34]. Recently, Sanchez-Perez et al. [35] introduced a wearable multimodal sensing system capable of capturing high-quality lung sounds in clinical settings, which can be instrumental in assessing cardiopulmonary health. Additionally, as addressed in Kraman et al. [30] the advancement of smart devices has significantly enhanced the utility of lung sounds. Over the next decade, we can anticipate substantial improvements in diagnostic capabilities, as emerging home smart devices are poised to transform how we manage and analyze respiratory health. These innovations will not only allow for continuous, real-time monitoring of lung sounds but also enable patients to address clinical questions and receive personalized feedback outside of traditional healthcare settings. This progress holds the promise of making early detection and management of respiratory conditions more accessible and efficient, potentially leading to better health outcomes and a more proactive approach to care.

In this study, we investigated the potential of NOC/BC for diagnosing pulmonary fibrosis. The findings suggest a promising direction for integrating such diagnostic tools into future smart devices, thereby enhancing their role in respiratory health management. Additionally, although the number of breath cycles in each lung sound file was audio-visually marked in Audacity by an experienced researcher in this study, we plan to integrate our proposed crackle detection system with our recently developed second derivative based automatic breathing phase detection technique [36]. This integration will enable a fully automated estimation of NOC/BC from recorded lung sounds.

Although, the automatic system showed good potential to differentiate subjects with radiological evidence of pulmonary fibrosis from subjects without, the study has several limitations: (i) the automatic system was tested on a small population and testing on a larger dataset will be needed for full validation, (ii) the automatic system counts all crackles (Velcro, fine, and coarse) but the threshold of ∼ 18 was established [23] for inspiratory Velcro crackles only, (iii) the physicians listened for Velcro crackles and classified subjects based on presence or absence of Velcro crackles only whereas the automated classifier decision is based on all types of crackles, (iv) the study used an existing threshold (∼18) [23] for classification; developing a method for automatic threshold selection based on lung sound characteristics is an area for further exploration, and (v) the performance of the IEM-FD filter [17] was not evaluated specifically on this dataset. Therefore, future research should focus on individually evaluating the different components of the automatic system on a larger and more diverse dataset which will test separately its ability to separate, verify, and count crackles.

## 7. Conclusions

Early diagnosis of fibrotic lung disease is important if emerging non-curative treatments are to be administered at an early stage of disease progression. It has been suggested that lung sounds, in particular Velcro crackles could be used as a non-invasive indicator of disease onset [14].

This study shows that an automatic system based on the average NOC/BC estimated from lung sounds recorded in the posterior lung bases can be used for diagnosis of pulmonary fibrosis. The classification performance of the automatic system was tested using a dataset containing equal numbers of fibrotic subjects (identified from pulmonary HRCT scans) and subjects with symptoms indicative of other pathological lung conditions.

In a comparison with two expert physicians asked to listen to the same lung sound recordings to identify the presence of Velcro crackles, the automatic system, had higher sensitivity (91.7 %) and similar specificity (59.3 %).

These preliminary results are very promising but need further validation with a much larger and more diverse dataset. Nevertheless, we contend that although radiological assessment must for now remain the gold standard for diagnosis of pulmonary fibrosis, a system such as described here has strong potential for diagnostic support, especially for assisting general practitioners to have confidence in their auscultatory assessment of lung sounds in these less commonly presenting diseases.

## Data Availability

All data produced in the present study are available upon reasonable request to the authors

## Acknowledgments

This work was supported by the NIHR Southampton Biomedical Research Centre, the Engineering and Physical Sciences Research Council (EPSRC), and the AAIR Charity.

